# Building process improvement capacity in epidemiologic research operations: The Nurses’ Health Studies experience

**DOI:** 10.1101/2025.11.20.25340660

**Authors:** Leanna Bassett, Vedika Vilas Patankar, Cizz-l N. Lockhart, Matt B. Mahoney, Janine Neville-Golden, Tanya L. Palmer, Nicole Romero, Jacqueline R. Starr

## Abstract

With declining infrastructure funding for large-scale epidemiologic cohort studies, continuous process improvement is essential to sustain research quality and impact. The authors recently undertook a Lean Six Sigma (LSS) training initiative with operational staff of the Nurses’ Health Studies, among the longest-running and most productive cohort studies, to modernize operations and build internal capacity for process improvement. Twenty staff members across operational domains completed a structured LSS curriculum and three improvement projects: (1) reducing median turnaround time for medical record retrieval by 90%, (2) decreasing the proportion undeliverable questionnaires by 31%, and (3) cutting biweekly freezer maintenance time by 43 person-minutes per cycle, yielding an estimated 6.5 person-weeks annually. Staff reported substantial increases in understanding and confidence in applying process improvement methods. Fourteen trainees completed all requirements for LSS green-belt certification. Though some participants questioned the need to learn a specialized framework, most valued the collaborative, data-driven approach. This experience demonstrates that LSS can be successfully adapted to non-clinical trial epidemiologic research, improving efficiency and data quality. The results support broader adoption of structured continuous improvement efforts to enhance the sustainability of epidemiologic research platforms.

## INTRODUCTION

Never has it seemed more important for epidemiologic research operations to achieve more with less. Even before recent shifts in federal research funding, the National Cancer Institute—and, to varying degrees, other NIH institutes—had already begun decreasing their commitments to provide infrastructure-specific funds for large-scale, longitudinal cohort operations. Stewardship of existing data is costly, and cohort operations must adapt to evolving technologies and research priorities. These exigencies make continuous process improvement imperative to maximize quality, efficiency, use of curated resources, and research impact.

The Nurses’ Health Studies (NHSs), ongoing since 1976 and collectively including over 300,000 participants, have played a pivotal role in shaping public health knowledge over several decades, contributing significantly to research on chronic diseases, women’s health, and epidemiology (1–3). This success is partly due to once-visionary large-scale tracking and data systems that use a custom computing language—innovations that anticipated now commonplace data, customer-relationship, and laboratory information management software. Five decades later, the system needs major restructuring. In planning data system modernization, we saw an opportunity to address undocumented processes, disorganized workflows, and bottlenecks in protocols.

Formal frameworks for quality and process improvement adapted from the manufacturing sector have been widely used to optimize healthcare delivery processes (4–6). Some frameworks have also been applied to FDA-regulated clinical trials research, of which Lean Six Sigma (LSS) is among the best recognized (7–12). We are unaware of any literature describing application to non-clinical trial epidemiologic research. We undertook LSS training of staff who operate the NHS cohorts. Here, we test the feasibility and usefulness of continuous improvement training in epidemiologic research through three process improvement projects.

## METHODS

### Overview of the NHSs and their operations

The first Nurses’ Health Study cohort (NHS) began in 1976 with national recruitment of over 120,000 nurses to be surveyed bi-annually by written questionnaire regarding diet, physical activity, disease diagnoses, and medications (3). A major focus has been on lifestyle factors contributing to risk of chronic disease and mortality. A second cohort, NHSII, added over 116,000 nurses beginning in 1989. It included a younger generation of women and a wider range of reproductive and lifestyle factors. Over time, protocols were implemented to collect a variety of biospecimens. Launched in 1996, the Growing Up Today Study (GUTS) included over 27,000 children of NHSII participants. A third nurse cohort, NHS3, began in 2010, incorporating male nurses, increasing racial and ethnic diversity, and transitioning to electronic-only data collection. NHS3 has included approximately 50,000 participants. Collectively, over 300,000 participants have been or continue to be followed through a large-scale research platform encompassing billions of data points and biospecimen samples. Use of these resources has led to the publication of nearly 5,000 peer-reviewed manuscripts and contributed to the implementation of national and international health policies.

### LSS in brief

LSS combines Lean Management, which reduces waste by eliminating non-value-added steps, and Six Sigma, which uses data-driven methods to improve quality and efficiency and reduce errors and variability. LSS offers a structured framework to assess baseline performance (e.g. cost, quality, and productivity), set improvement goals, and implement and sustain solutions.

A core feature of LSS is collaborative process mapping: LSS facilitators work with subject-matter experts (the staff who perform the work) to document each process step and its inputs, outputs, and key performance indicators. This mapping distinguishes value-added from non-value-added activities and helps identify sources of waste, such as delays, unnecessary movement, over-processing, errors, and underused staff skills.

Projects follow the DMAIC roadmap (13, 14). Define: clarify goals and identify critical-to-quality characteristics. Measure: Develop and validate measurement systems, then collect baseline data. Analyze: Use data and structured tools to identify root causes of inefficiency or defects. Improve: Design, test, and implement targeted solutions. Control: Establish monitoring and feedback systems to sustain gains and respond to future variation.

Many LSS tools will be familiar to epidemiologists, including Pareto charts and hypothesis tests (e.g. t-tests, ANOVA, or non-parametric two-sample tests). Other tools are specific types of charts or analysis based on simple descriptive statistics, tests, or reliability analysis, such as measurement systems analysis. LSS also employs process-specific tools less common in epidemiology (see below, under “LSS methods used by all the teams”)(15).

### LSS training program

We pursued LSS training for staff, most of whom had been in their roles for many years (described in Appendix 1). We identified training organizations through online searches and referrals and obtained external references for the company hired. We asked operations managers to shift work priorities to accommodate LSS training and projects.

The training company helped design a program for 20 staff members. The 40-hour classroom curriculum was divided into two 20-hour weeks, one in person and one by video conference in September 2024. We included staff from each area of operations: NHS3 and GUTS, Programming, Disease and Tissue Follow-up, Biorepository, NHS and NHSII Questionnaires and Coding, Bioinformatics, and Transformation (Appendix 1). The curriculum combined lectures with interactive activities, including hands-on work on trainees’ own projects, allowing teams to apply DMAIC tools under direct mentorship.

After the classroom curriculum, we surveyed participants (Appendix 2), using Likert-scale and two open-ended questions: What did you like most about the training? What did you like least about the training, and what are your suggestions for improvement?

In five teams, trainees then conducted projects under the guidance of the company’s “black belt” mentor. Every trainee was offered the opportunity to become LSS green belt certified, which would require documentation of their work, an oral final presentation about the process and results, and passing of a brief online exam. After the six-month project period, we surveyed participants again.

### Specific LSS projects in NHSs operations

Several months before training, the transformation project manager, who coordinated the effort, and the LSS mentor helped teams identify projects amenable to the six-month training period, which required collection of at least 30 baseline data points, development of an improvement plan, implementation of improvements, collection of at least 30 post-improvement data points, and development of a control plan to maintain improvements. We identified projects that would be of value though relatively simple. For example, we limited attention to processes in which most could be controlled directly by trainees, whereas many processes involve movement of materials or information between operational teams. After beginning their projects, several teams needed to iterate to identify projects that would meet the requirements.

We focus here on three projects with the most generalizable learnings. The disease follow-up team sought to reduce the turnaround for obtaining medical records documenting a breast cancer diagnosis. The NHS/NHSII questionnaire team sought to reduce the proportion of undeliverable questionnaires. The biorepository team sought to reduce the time required to perform biweekly freezer maintenance, a physical and time-consuming process in which over 100 liquid nitrogen freezers in two locations must be checked, filled, and cleaned.

### LSS methods used by all the teams

LSS training covers a variety of qualitative and quantitative tools. For certification, teams were required to demonstrate use of one set of tools and could choose from among others. Common to all teams were the SIPOC (suppliers, inputs, process steps, outputs, and “customer” requirements) chart (16), current state value stream mapping (17), swim lane mapping, a process input map (18), a cause-and-effect matrix (19), and failure mode and effects analysis (20).

The SIPOC provides a high-level map of process components, information that feeds into subsequent charts. Current state value stream mapping helps identify waste by tracking process time spent adding value, i.e., in operation, versus non-value-added time. Swim lane maps help visualize process steps, including decision points and dependencies across people or resources. The process input map is a more detailed version of the SIPOC, delineating inputs and outputs for each process step.

The cause-and-effect matrix and failure mode and effects analysis are semi-quantitative tools in which row entries are assigned scores to help rank their importance. The former helps identify the most important inputs influencing the most important outputs. In the latter, scores for the severity of a failure mode’s effect, how frequently it occurs, and how easily it is detected are combined into an overall risk priority number, focusing attention on elements requiring control to assure quality and efficiency.

### Statistical analysis of LSS project data

We performed graphical analysis by using Stata version 16.1 (StataCorp; College Station, TX) and LaTeX within Overleaf, including a process map (project 1), Kaplan-Meier curves (project 1), a Pareto chart (project 2), bar charts (projects 2 and 3), box plots (project 3), and a scatter plot (project 3). In addition to the qualitative and semi-quantitative analyses described above, we performed statistical analyses by using Stata version 16.1 (StataCorp; College Station, TX). Hypothesis testing to compare post-improvement to baseline measurements was done by testing differences in cycle times, accounting for censoring, in project 1 (log-rank test), differences in proportions in project 2 (z-test), and differences in means in project 3 (unpaired t-test).

## RESULTS

Twenty participants began the training, with 19 (95%) completing the classroom curriculum. One planned only to attend the classes, a second dropped out of training, and a third resigned. The remaining 17 (85%) participated in projects. The Director of Strategic Initiatives and the Transformation Project Manager designed the survey and were not considered survey respondents.

### Staff evaluation of training following classroom curriculum

Ten of 18 (56%) to whom we sent the training evaluation at the end of classroom training completed the survey. Reported good understanding of LSS and process improvement rose from two (20%) before training to seven (70%) after the classroom portion (Appendix 3). Five (50%) rated their experience as good at this point. Four (40%) found the material relevant to their work, six (60%) anticipated applying it to improve processes, and four (40%) believed it would help them professionally.

Qualitatively, the overwhelming response to what participants liked most was interacting in group activities, including didactic exercises and the projects. Participants liked least the perceived lack of applicability of training materials geared for manufacturing. Several participants felt they had not been adequately prepared and had not anticipated the time commitment. Some wondered about the relevance of Six Sigma to our work and whether Lean training alone would be more valuable. One respondent suggested, “Regular time and space dedicated to these conversations on improving specific projects and processes with multiple levels of staff without the Lean Six framework could be just as useful.”

### Staff evaluation of training after the six-month project period

Twelve of 15 participants (80%) completed the post-project training evaluation (Appendices 1 and 4). The percent reporting a good understanding of LSS and process improvement increased from 70% (7 of 10) to 92% (11 of 12). One (8%) rated the training experience as poor and six (50%) as average. Six (50%) deemed the training material relevant to their work, and nine (75%) believed they would be able to apply what they learned to improve processes. The percentage who believed the training would help them develop professionally increased from 40% (4 of 10) before the projects to 75% (9 of 12).

Multiple participants expressed appreciation that they saw immediate improvements and again acknowledged the importance and enjoyment of teamwork. One recognized the usefulness of the worksheets for problem-solving (despite their tediousness) and the importance of tracking process data. The least liked aspects of training were similar to those reported earlier.

### Project completion and LSS green belt certification

Four of five subprojects were successfully completed, i.e., meeting goals defined according to a written charter, with demonstrated work in each of the five DMAIC phases within six months. We omit description of one subproject relating to approval of new user accounts because it was not specific to epidemiology. A fifth subproject was not completed. This was partly because the programming group struggled to define a project with no process steps involving other groups and that could generate a sufficient number of data points to meet green belt certification requirements in six months. Fourteen trainees completed all training requirements and received LSS green belt certification.

### Project 1: Turnaround time for receipt of medical records reduced by 90%

For purposes of this project, a disease follow-up workflow cycle began upon receipt of a participant’s signed e-consent authorizing request of her medical record and ended upon closeout of the record (Figure 1A). Interim steps involved ascertaining providers’ contact information, preparing mailings, verifying completeness and quality of received records, scanning the record, and logging results. In the baseline period, the disease follow-up team had sent 336 breast cancer medical records requests in May and June of 2022, January 2023, and January 2024. Since then, 262 (78%) of the records had been received back, with a median turnaround time of 357 days. The team developed the goal of reducing the average turnaround time by 75%, to 90 days. In the define and measure phases, they identified three key process input variables: standard operating protocols (SOPs), operators, and e-consenting printing (Figure 1B). Sub-optimal SOPs and operator variation were also highlighted as potential causes of delays from failure mode and effects analysis in the analyze phase (not shown).

**Figure 1.**
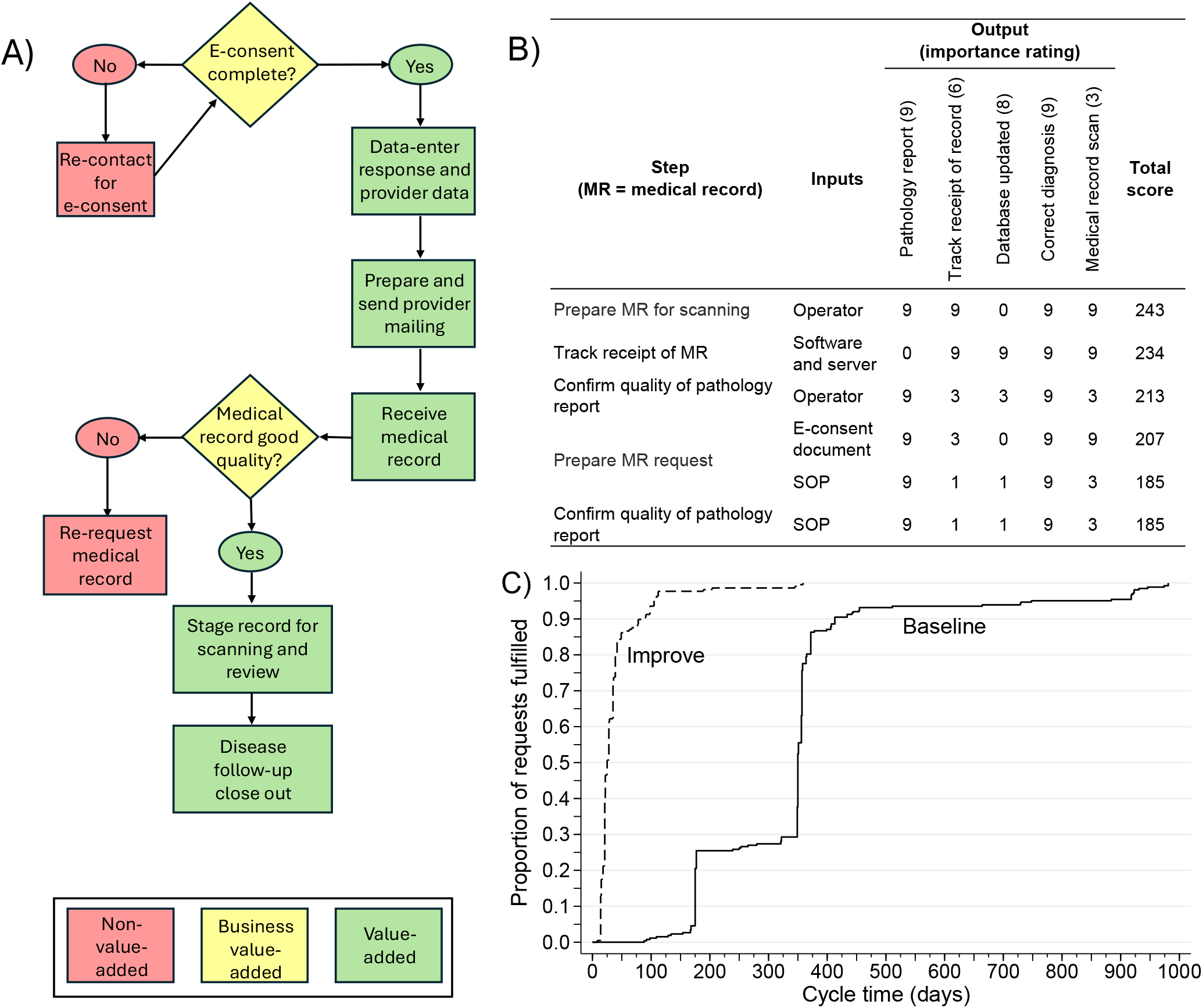
First of three projects undertaken by Nurses’ Health Studies operations staff as part of process improvement training from September 2024 to March 2025: the Disease and Tissue Follow-up group reduced the time to receive and track breast cancer medical records from participants’ providers. (A) A workflow cycle began upon receipt of a participant’s signed e-consent (first yellow diamond) and ended upon closeout of the record (final green rectangle). Interim steps involved ascertaining providers’ contact information, preparing mailings, verifying completeness and quality of received records, scanning the record, and logging results. (B) In cause-and-effect analysis, two of the top inputs having the greatest effect on the outputs were the operator (having effects in multiple process steps). In this analysis, outputs’ importance ratings could range from one to ten (from least to most important), and the importance of an input could be scored as 0, 1, 3, or 9 (for no effect or small, medium, or large effect, respectively). The total score is the sum of the inputs’ effect scores multiplied by each output’s importance score. SOP = Standard Operating Procedure. (C) Kaplan-Meier curves showing time from medical record request to receipt before and after implementation of process improvements. There were 336 baseline records and 206 records requested after implementation of improvements, with median cycle times of 357 and 35, respectively (*p* < 0.00005, Mann–Whitney). Curves account for right-censoring of unfulfilled requests.

The team tested the following improvements simultaneously: they modified the e-consent workflow to highlight a checking step, improved the mail room SOP, retrained staff, and added visual reminders regarding specific steps. Two hundred six new requests were sent in February 2025. By June 2025, 134 (65%) had been received, with a median turnaround of 35 days, a 90% reduction from baseline (log-rank *P*=3.16×10^−124^)(Figure 1C).

In the control phase, in addition to implementation of updated SOPs and continuing use of visual cues, managers will track process variability quarterly, using the reports developed during the project.

### Project 2: Proportion of undeliverable questionnaires in NHS and NHSII reduced by 31%

The NHS and NHSII questionnaire and coding team randomly selected 546 participants for whom questionnaires were mailed from January to October 2024, with 345 (63.2%) questionnaires undeliverable. In the define and measure phases, the team targeted a ten percent reduction in the proportion of returned questionnaires. Current state value stream mapping showed that the vast majority of cycle time was non-value added, due to delays (not shown), and the SIPOC chart indicated updating of participants’ contact information as the input impeding successful delivery of questionnaires (Figure 2A). The majority of postal error codes were coded as “not delivered as addressed,” and the next largest proportion had “no forwarding address” or “insufficient address” (Figure 2B). In the measure phase, other factors were shown to be more informative than the postal error code. Qualitative analysis, including failure mode and effects analysis, indicated that research assistants were expected to update participant files only when they had extra time after completing other tasks, resulting in failure to update files and repetitive returns of undeliverable questionnaires (Figure 2C). Control charts by subgroups indicated that the proportion undeliverable depended on the cohort and age group (not shown).

**Figure 2.**
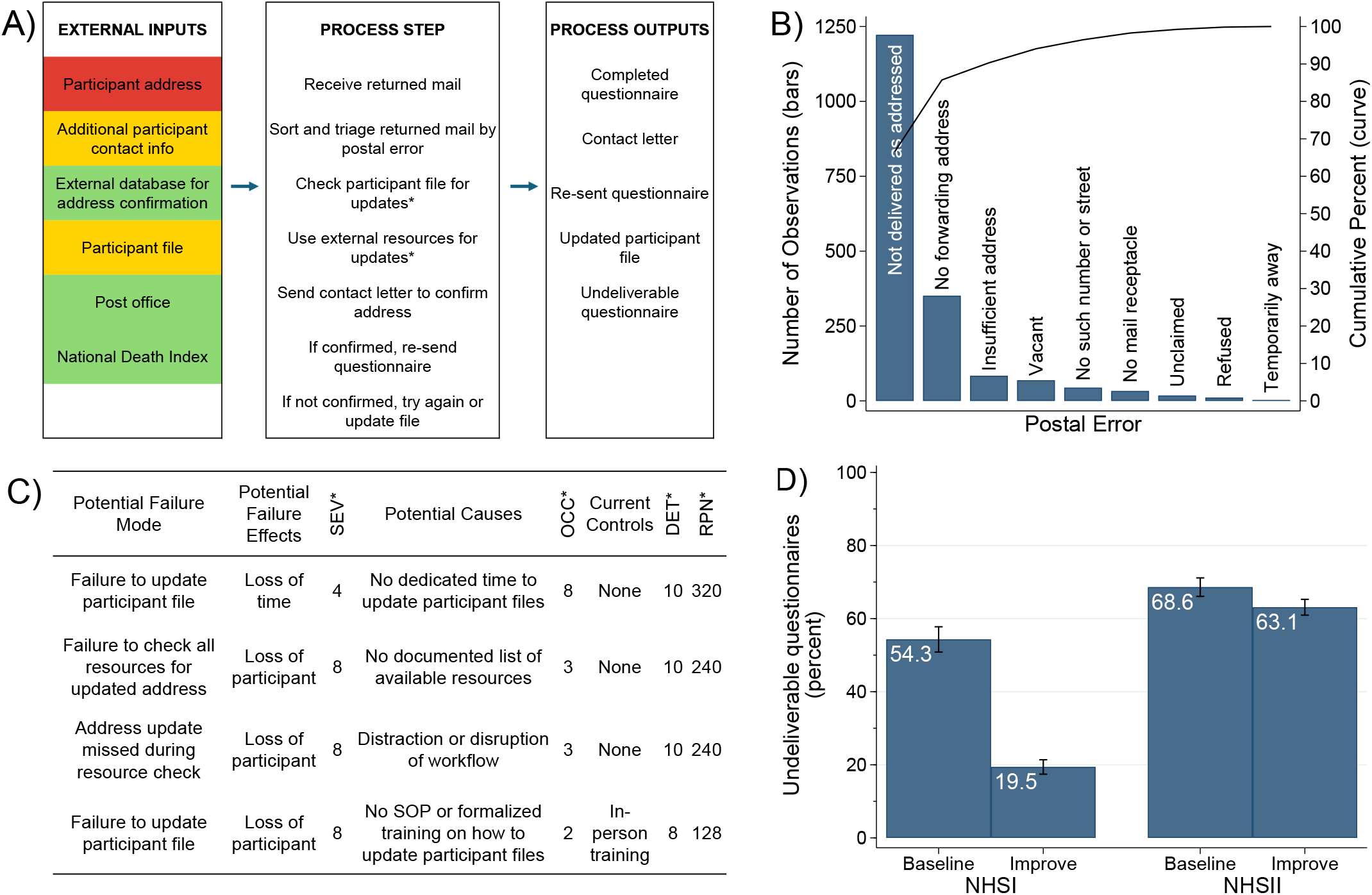
Second of three projects undertaken by Nurses’ Health Studies’ operations staff as part of process improvement training from September 2024 to March 2025: the Questionnaires and Coding group reduced the proportion of questions returned as undeliverable. (A) The SIPOC (suppliers, inputs, process step, outputs, customer requirements) chart showed that participants’ addresses, contact information, and other information in their files were inputs with negative effects on outputs (red=known negative effects, yellow=believed negative effects, and green=no negative effects). Note that we omitted the suppliers and customer requirements from this chart. (B) Pareto chart shows the distribution of postal errors causing questionnaires to be returned undelivered (bars) and cumulative percent over the error categories ordered from largest to smallest (curve). Two thirds had no specific reason for the questionnaires to be undelivered. The most common specific reason was lack of a forwarding address, and the next most common reason was an insufficient address. (C) Excerpt from the team’s failure mode and effects analysis (FMEA), showing the four highest-risk modes of failure. *SEV=severity, measuring the potential impact of a failure (range 1-10; higher scores indicate greater impact). *OCC=occurrence, or likelihood or frequency of a failure type (range 1-10; higher scores indicate greater probability or frequency of failing through that mode). *DET=detection, or how easily a failure is detected (range 1-10; higher scores indicate greater difficulty in detecting a failure of this type). *RPN=risk priority number, calculated by multiplying the SEV, OCC, and DET scores (range 1-1000, and higher scores indicate higher risks associated with a failure type). (D) Comparing the improvement to the baseline phase, the percentage of questionnaires returned as undeliverable dropped from 54.3% to 19.5% in the NHS (*P* = 0.00005), from 68.6% to 63.1% in NHSII (*P* = 0.1), and from 63.2% to 43.6% overall (*p* < 0.00005).

The team made improvements by revising the SOP to allot dedicated time to updating of participant files and to prioritize returned mail by participant information rather than by postal error codes. They also improved documentation and tracking of undeliverable questionnaires. In the improve phase, they randomly selected a different 901 participants whose questionnaires had been sent during the same January to October 2024 period. With 393 of these undeliverable, the improvements resulted in a 31% reduction in the percent of undeliverable questionnaires, from 63.2% to 43.6% (p<0.00005; Figure 2D). To maintain improvements, the team will generate similar charts on a quarterly basis.

### Project 3: Time spent on biweekly freezer maintenance reduced by 6.5 person-weeks per year

From late October 2024 to early February 2025, the biorepository team collected data on 53 baseline freezer maintenance cycles across two locations, averaging 236 person-minutes per cycle. In the define phase, they conservatively targeted a 10% reduction in cycle times. In the measure phase, value stream mapping indicated that over 20% of time was on non-value-added transportation time (walking between freezer room locations)(Figure 3A). This and other analyses helped identify critical inputs affecting cycle times: SOPs, tools and equipment, travel time, and documentation. In the analyze phase, failure mode and effects analysis highlighted SOPs and staff training and retraining as vulnerabilities (not shown). Graphical analysis suggested that having five or six staff unnecessarily increased cycle person-time compared with smaller numbers of staff (Figure 3B).

**Figure 3.**
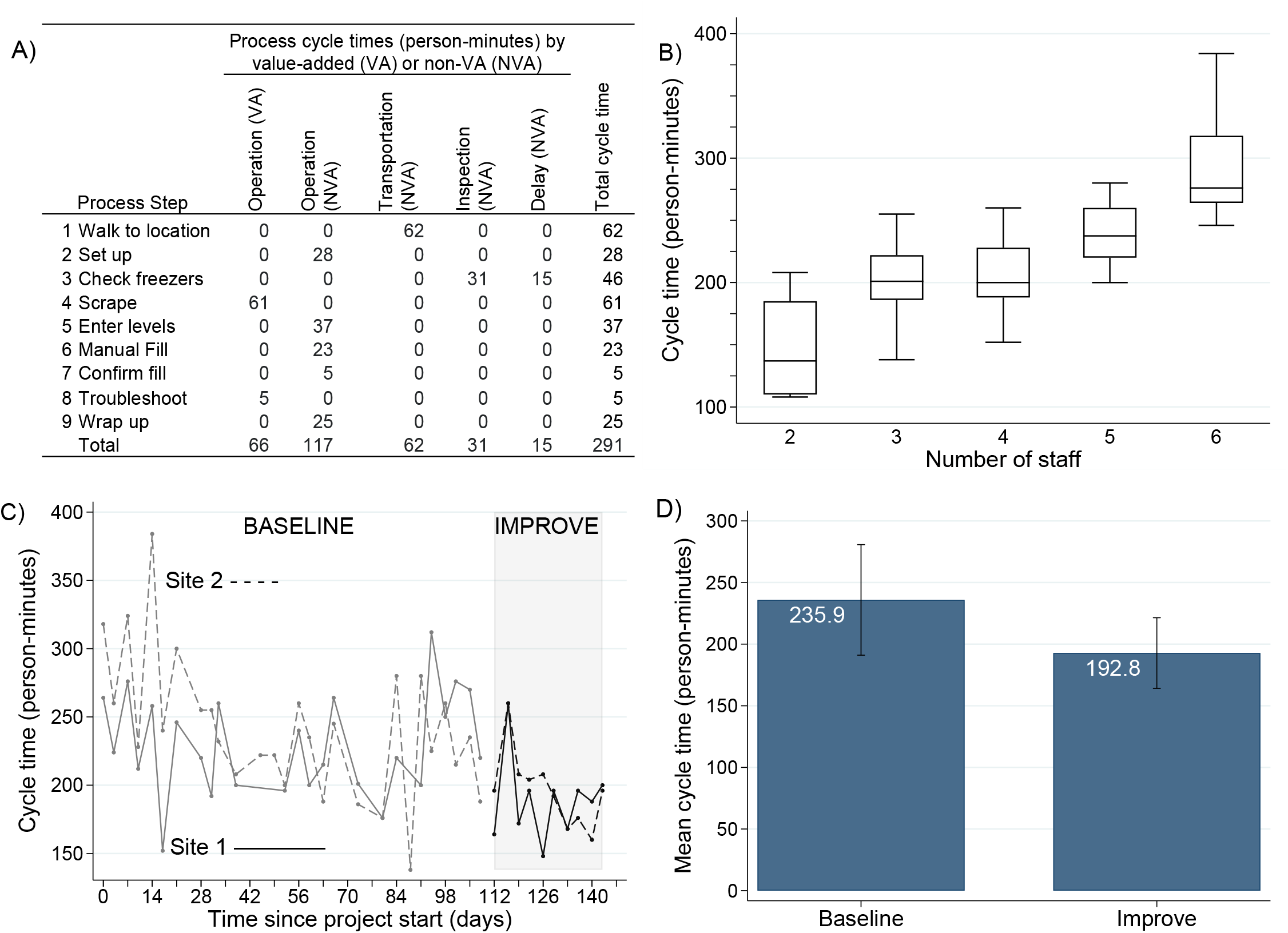
Third of three projects undertaken by Nurses’ Health Studies’ operations staff as part of process improvement training from September 2024 to March 2025: Biorepository staff reduced biweekly cycle time for checking and maintaining liquid nitrogen tank freezers. (A) In current state value stream mapping, process steps are classified according to whether they add value or not and, for non-value-added steps, the type of waste. Twenty-three percent of time was spent in operations that add value. (B) Graphical analysis showed that, in the baseline period, mean cycle times increased with the number of staff working freezer check cycles. (C) Charting cycle times by location (solid line, site 1; dashed line, site 2) showed longer times and greater variability in the baseline period (grey curves and unshaded plot) than after improvements were implemented (black curves and shaded plot). (D) Mean cycle time dropped from 235.9 person-hours (±44.9 SD) at baseline to 192.8 person-hours (±28.7 SD) post-improvement (*P* = 0.0002, unpaired t-test).

Improvements included revising the SOP—capping the number of staff at four and reducing checking of modern, efficient freezers to once weekly; updating tools, purchasing a greater number and placing them near each freezer; identifying the fastest travel route; and retraining staff. Improvements (n=20 cycles) yielded an 18% reduction in mean cycle time to 193 person-minutes (*P*=0.0002), with reduced process variation (Figures 3C and D). Multiplying by four weekly cycles and using benchmarks for standard work weeks, we estimated the time savings would translate to a 6.5-week reduction in staff-time annually.

To sustain these improvements, visual cues were placed in the freezer rooms, on freezer room maps, and in the SOP. The control plan also included weekly freezer spot checks, biweekly check-ins with staff, quarterly cycle time monitoring, and enhanced managerial oversight. An outside consultant was hired to teach staff more about freezer maintenance, reducing reliance on managers.

## DISCUSSION

The core data infrastructure of the NHSs has changed little since its inception. Historically, workflows prioritized quality over operational efficiency. Dependence on grant funding, a sense of urgency to produce results, and a drive to improve public health instilled a culture of being “too busy to stop to sharpen the saw.” Whether voiced or not, this mindset is common across large-scale epidemiologic research. Early in our transformation process, staff identified many inefficiencies they wanted to improve. Barriers to direct action included heavy workloads, risk of burnout, and limited opportunities for skills development. We believe making time for process improvement is a solution, not a contributor, to these challenges.

The immediate goal of the training described here was to build internal capacity for continuous process improvement. The overarching goal was to enhance sustainability of the cohort infrastructure. Based on only a six-month experience, this report provides evidence of early gains rather than proof of durable process-improvement capacity. Demonstration of sustained capacity would require a longer time horizon (21–23). Nevertheless, such longer-term capability depends on short-term gains, and the survey responses and subproject results are early indicators of progress towards both building capacity and improving infrastructure efficacy (21–26). With staff trained across operations, the emerging capacity reflected in these early results provides a foundation for broader implementation.

Over 90% of trainees reported a good understanding of LSS and process improvement after the training, compared with 20% before the training. By the end of the project period, 75% believed they could apply what they learned to improve processes, and 75% expected the training would help them professionally. The survey response rate improved after the project period, possibly because this was a more selected group. We do not claim the results are generalizable and may have been more negative had non-respondents felt less favorable than respondents. Nevertheless, continuous process improvement does not require that all staff members be equally trained or equally receptive to the curriculum and framework (21, 22).

All three subprojects yielded time-saving improvements that will reduce study overhead while strengthening core epidemiologic functions. Decreasing the proportion of undeliverable questionnaires reduces loss to follow-up and non-response bias. Accelerating medical record retrieval enables more timely and complete diagnostic ascertainment and reduces the potential for disease misclassification. Improvements to freezer-maintenance workflows help preserve specimen integrity and minimize measurement error and missing data (due to lost samples). Together, these operational gains enhance data completeness and validity, thereby mitigating threats to internal validity.

One of the challenges we faced is apparent from the evaluations of the classroom curriculum, which emphasized examples from manufacturing. We later learned that some LSS trainers specialize in transactional processes, the LSS term for service-oriented processes. It is well-documented that acceptability of change frameworks is strongly influenced by how well they align with staff roles and work tasks (25, 27, 28). Better informing the trainer(s) about our work ahead of time may have improved training effectiveness and trainees’ engagement and satisfaction.

In preparing for training, we learned that no amount of pre-training communication with staff appears to be too much. The training company helped groups develop their project proposals and provided a project roadmap before class began. Internally, we sent repeated emails, held multiple meetings with each group, and worked with managers to set realistic expectations. Despite this effort, some participants underestimated the required time commitment. Their frustration was exacerbated by the paperwork and documentation required for certification (for those who chose to pursue it). Yet, as acknowledged by one survey respondent, even the documentation steps were part of the learning, and the documentation facilitates sharing of results and narratives with other stakeholders. These reactions are consistent with implementation research showing that perceived burden and effort are key determinants of acceptability and uptake, particularly in knowledge-work settings (21, 29, 30).

Though the LSS framework may strike some as commonsensical and not worth the cost of training, trainees’ understanding of process improvement increased greatly, and an internally led training program would also have been costly to organize. LSS is often implemented company- or institution-wide, with designated staff trained to the green belt level and others introduced to the basic principles. Our approach—training a large proportion of subject-matter experts to the green belt level—was less typical but appropriate in this context: with few redundancies, nearly every staff member occupies a role that could be considered critical. This approach was also motivated by a desire to provide career and skills development opportunities as inclusively and equitably as possible and to foster a broad culture of process improvement, a known facilitator of long-term acceptability and engagement.

As with many organizational change initiatives, staff varied in readiness, interest, and perceived fit of the framework with their day-to-day work. Implementation research emphasizes that acceptability—shaped by perceived relevance, clarity of expectations, and the cognitive or time burden associated with new practices—plays a critical role in achieving durable improvement (23, 29, 30). These dynamics were evident: staff welcomed opportunities for collaborative, structured problem-solving and saw value in improving long-standing workflows, yet some aspects of the curriculum and documentation requirements felt less aligned with their roles or prior experience. Recognizing this variation as expected rather than exceptional helps clarify why adaptation of process-change frameworks to research-operations environments is essential for long-term sustainability (29, 30).

A related challenge, with implications for long-term sustainability, concerned the balance between Lean and Six Sigma content. Several staff felt only Lean methods were directly applicable and that collecting new process data was not worthwhile. In all three projects, however, quantitative statistical analysis ultimately yielded some of the most unexpected and useful insights. For example, staff had believed postal error codes to be the primary predictor of undeliverable questionnaires, and data about participants themselves proved more informative. Biorepository managers thought that including the entire staff in freezer maintenance saved time by sharing work; graphical and statistical analysis showed clear limits to these gains, with excess travel time between sites outweighing the benefit of involving more than four staff members.

LSS and similar frameworks are sometimes criticized for the potential to dehumanize work or cause layoffs (31–33). Our goal was not staff reduction but to reduce the risk of burnout, to enable research platform expansion, and to increase productivity and quality within fixed costs. Initiating training in process improvement is one of many steps we are taking as part of broader transformation.

Despite initial reservations about LSS’s applicability to epidemiologic research, our experience demonstrates that, with thoughtful adaptation and inclusive engagement, such frameworks can yield substantial operational benefit—we cut costs and improved data quality—and are worthy of broader adoption in the field. Though there may be a few already highly efficient outliers, many epidemiologic infrastructure groups could benefit if they briefly stop sawing to sharpen the saw.

## Data Availability

All data produced in the present study are available upon reasonable request to the authors.

