## Supplemental tables and information for "Building process improvement capacity in epidemiologic research operations: The Nurses’ Health Studies experience"

**Supplemental tables and information for:**
**Building process improvement capacity in epidemiologic research operations:**
**The Nurses' Health Studies experience**

**Authors:** Leanna Bassett[1], Vedika Vilas Patankar[2], Cizz-l N. Lockhart[2], Matt B. Mahoney[2], Janine Neville-Golden[2], Tanya L. Palmer[2], Nicole Romero[2], Jacqueline R. Starr[2]

**Affiliations and contact information:** [1]Research Operations, Brigham and Women's Hospital, Boston, Massachusetts, United States; [2]Channing Division of Network Medicine, Department of Medicine, Brigham and Women's Hospital and Harvard Medical School, Boston, Massachusetts, United States

**List of Appendices**

**Appendix 1.** Selection and description of Lean Six Sigma training participants among Nurses' Health Studies staff, including tenure and attrition.

**Appendix 2.** Survey questions administered to Nurses' Health Studies operations staff after a 40-hour Lean Six Sigma curriculum and, again, after a six-month project period.

**Appendix 3.** Participant evaluations after a 40-hour Lean Six Sigma training curriculum was given to Nurses' Health Studies operations staff.

**Appendix 4.** Participant evaluations after a six-month Lean Six Sigma project period during which Nurses' Health Studies operations staff were mentored by black belt mentors.

**Appendix 1. Selection and description of Lean Six Sigma training participants among Nurses' Health Studies staff, including tenure and attrition.**

**Participant selection and description**

We focused selection of training participants on operational staff and not faculty. Although faculty oversee cohort operations and could benefit from process improvement training, they are generally not involved in day-to-day workflows, and most have access to discretionary resources for external training. The sole exception was the Director of Strategic Initiatives, who leads the Nurses' Health Studies (NHSs) transformation effort and whose role is directly relevant to operational modernization.

A staff inclusion criterion was holding a role relevant to core cohort infrastructure, i.e. not employed solely to support an ancillary study or individual faculty member. We applied several exclusion criteria. We excluded individuals who 1) held administrative roles not directly tied to data collection, data management, or the biorepository; 2) were *per diem* or temporary employees, including part-time student research assistants; or 3) had been hired within the prior year. Although newer employees bring valuable perspectives, we sought to train individuals who would be positioned to sustain a process-improvement culture and to mentor colleagues over time. Thus, we prioritized long-tenured staff and operational managers while also including junior staff with unique or specialized roles.

The Transformation Project Manager conferred with operations managers, faculty principal investigators, and members of the Transformation Management Committee to develop a list of staff to invite. Fewer than five individuals declined participation due to scheduling conflicts (e.g., planned time off or caregiving responsibilities).

Across operational categories, we selected 17 of 32 (53%) staff meeting inclusion criteria:

- Data collection: 8 of 10 (80%)
- Programming (coding): 6 of 15 (40%)
- Biorepository: 3 of 7 (43%)

The selected proportion was higher among data-collection staff because workflows and responsibilities vary substantially across the four cohorts, making these roles less interchangeable.

Three additional trainees were included:

1. the Director of Strategic Initiatives (described above),
2. the Transformation Project Manager, who coordinated the training effort and other transformation tasks, and
3. a divisional bioinformatics project manager who was supporting migration of the NHSs biorepository from a legacy system to a modern laboratory information management system (LIMS).

**Participants' tenure and attrition**

The majority of training participants had been in their roles, or closely related roles, for more than ten years, and several for much longer. One programmer notified us at the outset that she would be unable to attend the full training, and one data-collection trainee withdrew during the classroom curriculum; both had long tenures in their roles. A third participant, one of the most junior and recent hires, left her position during the project phase of the training.

The Director of Strategic Initiatives and the Transformation Project Manager facilitated the organization and delivery of the curriculum and were not considered potential survey respondents. Of the remaining 18 trainees, ten (56%) completed the post-classroom curriculum survey. We administered the second, post-project survey only to the 15 trainees who participated in the projects—regardless of project completion, again excluding the Director of Strategic Initiatives and Transformation Project Manager. Twelve (80%) responded. We could not assess potential non-response patterns for either survey because no questions about tenure or roles had been asked, to maintain anonymity and keep the surveys brief.

**Appendix 2.** Survey administered to Nurses' Health Studies operations staff after a 40-hour Lean Six Sigma curriculum and, again, after a six-month project.

| Question (survey two changes are noted in parentheses in italics) | Possible responses |
|---|---|
| 1. How would you rate your overall training experience? | Very poor, poor, average, good, excellent |
| 2. The training material was relevant to my role/position. | Strongly disagree, Disagree, Neutral, Agree, Strongly agree |
| 3. I will be able to apply what I have learned in this training and use it to improve processes within my area. | Strongly disagree, Disagree, Neutral, Agree, Strongly agree |
| 4. This training will help me develop professionally. | Strongly disagree, Disagree, Neutral, Agree, Strongly agree |
| 5. I have (*had*) the information and materials necessary to achieve Green Belt certification. | Strongly disagree, Disagree, Neutral, Agree, Strongly agree |
| 6. I have (*had*) adequate support in my division to achieve Green Belt certification. | Strongly disagree, Disagree, Neutral, Agree, Strongly agree |
| 7. I have adequate support in division to continue process improvements beyond this training and certification. | Strongly disagree, Disagree, Neutral, Agree, Strongly agree |
| 8. I would recommend this training be offered in my division again in the future. | Strongly disagree, Disagree, Neutral, Agree, Strongly agree |
| 9. I would recommend this training to other research groups within the institution. | Strongly disagree, Disagree, Neutral, Agree, Strongly agree |
| 10. (*In the first survey only*) How would you describe your understanding of lean six sigma and process improvement prior to the start of this training? | Limited, moderate, good, strong, expert |
| 11. (*10. In the second survey*) How would you describe your understanding of lean six sigma and process improvement after this training? | Limited, moderate, good, strong, expert |
| 12. (*11. In the second survey*) What did you like most about the training? Please be specific. | |
| 13. (*12. In the second survey*) What did you like least about the training and what are your suggestions for improvement? Please be specific and constructive. | |

**Appendix 3.** Participant evaluations after a 40-hour Lean Six Sigma training curriculum was given to Nurses' Health Studies operations staff.

| **Question (abbreviated)*** | **Number (percent) choosing each response options (N=10)**** | | | | | **Mean response** |
|---|---|---|---|---|---|---|
| | *Very poor* | *Poor* | *Average* | *Good* | *Excellent* | |
| 1. Training experience | 1 (10.0) | 3 (30.0) | 1 (10.0) | 4 (40.0) | 1 (10.0) | 3.1 |
| | *Strongly disagree* | *Disagree* | *Neutral* | *Agree* | *Strongly agree* | |
| 2. Training material relevant | 2 (20.0) | 2 (20.0) | 2 (20.0) | 1 (10.0) | 3 (30.0) | 3.1 |
| 3. Will be able to apply learning | 1 (10.0) | 1 (10.0) | 2 (20.0) | 4 (40.0) | 2 (20.0) | 3.5 |
| 4. Will help me develop professionally | 1 (10.0) | 1 (10.0) | 4 (40.0) | 0 (0.0) | 4 (40.0) | 3.5 |
| 5. Have resources to achieve green belt certification | 1 (10.0) | 1 (10.0) | 3 (30.0) | 5 (50.0) | 0 (0.0) | 3.2 |
| 6. Have support to achieve green belt certification | 0 (0.0) | 0 (0.0) | 2 (20.0) | 8 (80.0) | 0 (0.0) | 3.8 |
| 7. Have support to continue process improvements | 1 (10.0) | 1 (10.0) | 3 (30.0) | 5 (50.0) | 0 (0.0) | 3.2 |
| 8. Recommend training in our division in future | 3 (30.0) | 2 (20.0) | 1 (10.0) | 3 (30.0) | 1 (10.0) | 2.7 |
| 9. Recommend training in our hospital in future | 2 (20.0) | 3 (30.0) | 1 (10.0) | 4 (40.0) | 0 (0.0) | 2.7 |
| | *Limited* | *Moderate* | *Good* | *Strong* | *Expert* | |
| 10. Understanding prior to training | 6 (60.0) | 2 (20.0) | 1 (10.0) | 1 (10.0) | 0 (0.0) | 1.7 |
| 11. Understanding after this part of training | 0 (0.0) | 3 (30.0) | 3 (30.0) | 4 (40.0) | 0 (0.0) | 3.1 |

*See Appendix 2 for the complete questions.

**Categories were scored 1, 2, 3, 4, or 5, from left to right for each set of possible responses.

**Appendix 4.** Participant evaluations after a six-month Lean Six Sigma project period during which Nurses' Health Studies operations staff were mentored by black belt mentors.

| Question (abbreviated)* | Number (percent) choosing each response options (N=12)** | | | | | Mean response |
|---|---|---|---|---|---|---|
| | *Very poor* | *Poor* | *Average* | *Good* | *Excellent* | |
| 1. Training experience | 0 (0.0) | 1 (8.3) | 6 (50.0) | 4 (33.3) | 1 (8.3) | 3.4 |
| | *Strongly disagree* | *Disagree* | *Neutral* | *Agree* | *Strongly agree* | |
| 2. Training material relevant | 1 (8.3) | 2 (16.7) | 3 (25.0) | 4 (33.3) | 2 (16.7) | 3.3 |
| 3. Will be able to apply learning | 0 (0.0) | 0 (0.0) | 3 (25.0) | 5 (41.7) | 4 (33.3) | 4.1 |
| 4. Will help me develop professionally | 0 (0.0) | 0 (0.0) | 3 (25.0) | 4 (33.3) | 5 (41.7) | 4.2 |
| Had resources to achieve green belt certification | 0 (0.0) | 1 (8.3) | 1 (8.3) | 7 (58.3) | 2 (16.7) | 3.9 |
| Had support to achieve green belt certification | 0 (0.0) | 1 (8.3) | 1 (8.3) | 2 (16.7) | 7 (58.3) | 4.4 |
| 7. Have support to continue process improvements | 0 (0.0) | 1 (8.3) | 2 (16.7) | 5 (41.7) | 4 (33.3) | 4.0 |
| 8. Recommend training in our division in future | 0 (0.0) | 3 (25.0) | 2 (16.7) | 4 (33.3) | 3 (25.0) | 3.6 |
| 9. Recommend training in our hospital in future | 0 (0.0) | 1 (8.3) | 4 (33.3) | 6 (50.0) | 1 (8.3) | 3.6 |
| | *Limited* | *Moderate* | *Good* | *Strong* | *Expert* | |
| 10. Understanding after this part of training | 0 (0.0) | 1 (8.3) | 6 (50.0) | 5 (41.7) | 0 (0.0) | 3.3 |

*See Appendix 2 for the complete questions.

**Categories were scored 1, 2, 3, 4, or 5, from left to right for each set of possible responses.
